# The maladaptive vascular response in COVID-19 acute respiratory distress syndrome and recovery

**DOI:** 10.1101/2021.05.20.21257542

**Authors:** David R. Price, Elisa Benedetti, Katherine L. Hoffman, Luis Gomez-Escobar, Sergio Alvarez-Mulett, Allyson Capili, Hina Sarwath, Christopher N. Parkhurst, Elyse Lafond, Karissa Weidman, Arjun Ravishankar, Jin Gyu Cheong, Richa Batra, Mustafa Büyüközkan, Kelsey Chetnik, Imaani Easthausen, Edward J. Schenck, Alexandra C. Racanelli, Hasina Outtz Reed, Jeffrey C. Laurence, Steven Zvi Josefowicz, Lindsay Lief, Mary E. Choi, Shahin Rafii, Frank Schmidt, Alain C. Borczuk, Jan Krumsiek, Augustine M.K. Choi

**Author notes:** Shared first authorship (David R. Price, Elisa Benedetti). Corresponding authors Corresponding Authors: Jan Krumsiek, 1305 York Avenue, New York, NY 10021, Augustin M.K. Choi, 1300 York Avenue, New York, NY 10021.

## Abstract

Vascular injury is a menacing element of acute respiratory distress syndrome (ARDS) pathogenesis. To better understand the role of vascular injury in COVID-19 ARDS, we used lung autopsy immunohistochemistry and blood proteomics from COVID-19 subjects at distinct timepoints in disease pathogenesis, including a hospitalized cohort at risk of ARDS development (“*at risk”*, N=59), an intensive care unit cohort with ARDS (*“ARDS*”, N=31), and a cohort recovering from ARDS (“*recovery*”, N=12). COVID-19 ARDS lung autopsy tissue revealed an association between vascular injury and platelet-rich microthrombi. This link guided the derivation of a protein signature in the *at risk* cohort characterized by lower expression of vascular proteins in subjects who died, an early signal of vascular limitation termed the *maladaptive vascular response*. These findings were replicated in COVID-19 ARDS subjects, as well as when bacterial and influenza ARDS patients (N=29) were considered, hinting at a common final pathway of vascular injury that is more disease (ARDS) then cause (COVID-19) specific, and may be related to vascular cell death. Among *recovery* subjects, our vascular signature identified patients with good functional recovery one year later. This vascular injury signature could be used to identify ARDS patients most likely to benefit from vascular targeted therapies.

## INTRODUCTION

Vascular injury has been linked to COVID-19 acute respiratory distress syndrome (ARDS) (1, 2), including the vascular complications of inflammation and thrombosis. Consistent with this, COVID-19 induced injury to the vascular compartment has been associated with complement activation and micro-thrombosis (3–5), systemic thrombosis (4, 6), and to dysregulated immune responses (7–9). However, this focus on inflammation and thrombosis limits our insights into other disruptions associated with aberrant vascular activation. In mice, endothelial overexpression of the angiocrine factor angiopoietin-2 (ANGPT2) induces vascular leakage and disrupts capillary-associated endothelial/pericyte interactions (10). These vascular alterations are countered by ANGPT2 neutralizing antibodies or platelet derived pericyte chemokines such as angiopoietin-1 (ANGPT1) and platelet derived growth factor B (PDGFB), demonstrating the homeostatic potential of circulating vascular proteins (10). This is phenocopied in humans where an elevated plasma ANGPT2 to ANGPT1 ratio has been linked to acute lung injury mortality (11). In the right context, ANGPT2 can also disrupt vascular angiogenesis and shorten vascular cell survival, specifically when vascular growth factors are limited (12). More precise understanding of ANGPT2 associated vascular disruptions in COVID-19 ARDS and ARDS generally may inform the timing and patient selection for targeted vascular therapies in ARDS going forward.

Heterogeneity in the vascular response to injury between patients may also be associated with COVID-19 ARDS outcomes. Vascular biomarkers have been linked to COVID-19 ARDS severity, including ANGPT2 (13) and vascular cell death is an increasingly recognized consequence of ARDS inflammatory signaling (2, 14–16). Caspase-mediated apoptotic endothelial cell death has been demonstrated in COVID-19 ARDS autopsies (2). However, the clinical significance of circulating vascular and cell death proteins, including their link to COVID-19 ARDS disease and recovery, remains unclear. We hypothesized that differences in circulating vascular proteins are associated with ANGPT2 and could have predictive ability throughout the natural history of COVID-19 ARDS. To test this hypothesis, we quantified vascular proteins first in lung tissue of COVID-19 autopsy patients and then in blood of COVID-19 subjects from distinct disease timepoints (early hospitalization, intensive care, and recovery) and linked this vascular injury signature to relevant clinical outcomes, including mortality.

## RESULTS

### Study cohorts represent COVID-19 ARDS from distinct disease timepoints

An *autopsy* cohort of 20 COVID-19 subjects was first used to evaluate vascular proteins in human lung tissue. We then analyzed blood vascular proteins in three COVID-19 cohorts at distinct disease timepoints: in early hospitalization before ARDS onset (*at risk* cohort), after ARDS onset (*ARDS* cohort), and after discharge from the intensive care unit (*recovery* cohort). A graphical description of the COVID-19 subjects, including blood sampling, intubation time, and death is shown in **Figure 1**. Baseline characteristics of these subjects are listed in **Supplementary Table 1**.

**Figure 1:**
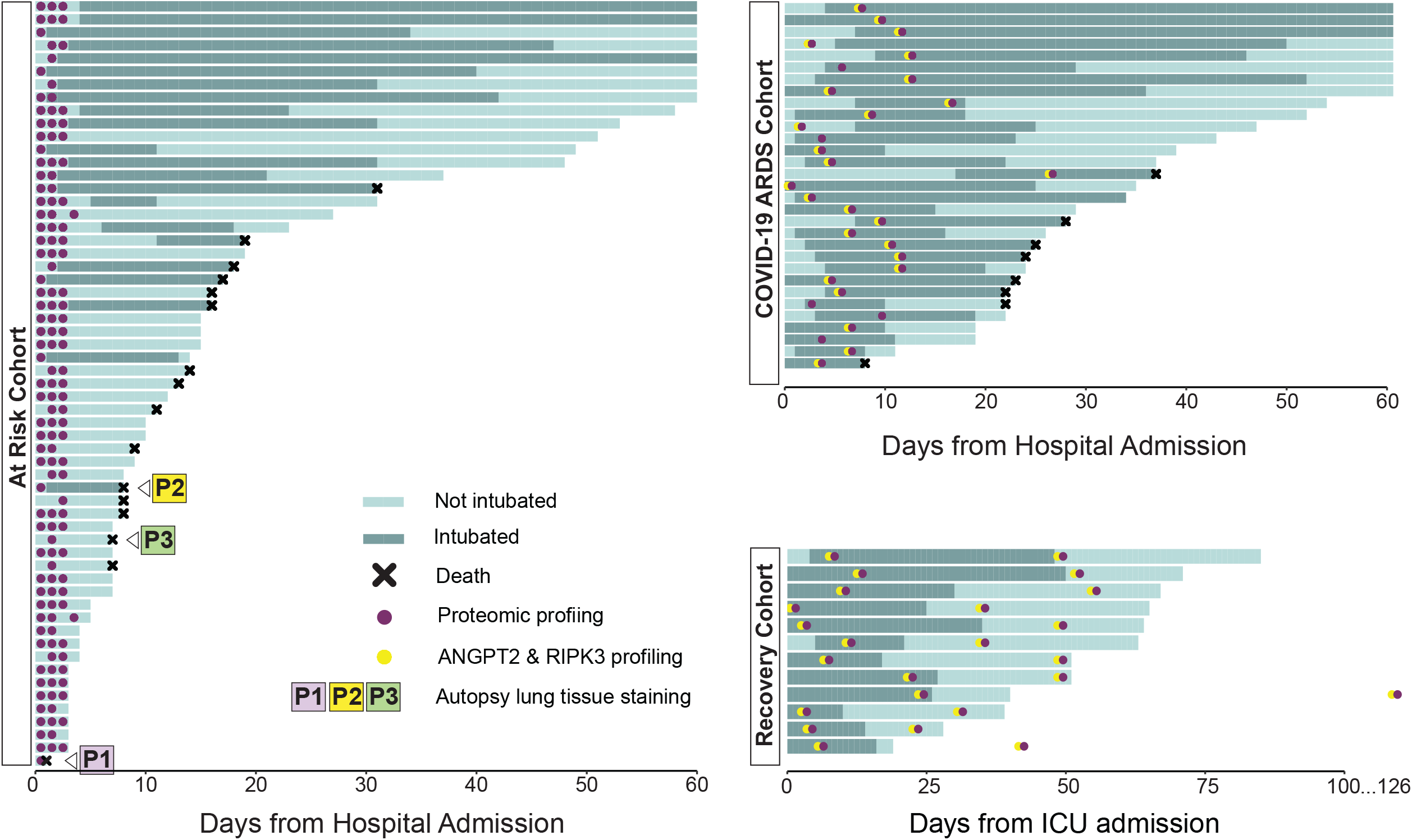
Overview of COVID-19 subjects in the *at risk* (N=59), *ARDS* (N=31) and *recovery* (N=12) cohorts. Each horizontal line corresponds to one individual. Subjects in the *at risk* cohort were sampled for proteomics between 1 and 3 times (purple dots). For three of the patients in this cohorts (P1, P2, P3) autopsy lung tissue staining was available. Subjects in the *ARDS* cohort were sampled once, within 10 days of ICU admission. *Recovery* subjects were sampled twice, once during their ICU stay and once after discharge from the ICU (median 31 days). Patients in the *ARDS* cohort were additionally profiled for ANGPT2 and RIPK3 (yellow dots) while *recovery* cohort subjects were profiled for ANGPT2 (yellow dots).

The *at risk* cohort included 59 COVID-19 subjects admitted to the medical floors of New York Presbyterian Weill Cornell Medical Center (WCM) that did not meet ARDS criteria at study enrollment. The median age of the *at risk* cohort was 69 years old and was majority male (64% male versus 36% female). Fifty-three percent of the cohort had hypertension and 15% had cancer.

The *ARDS* cohort included 31 COVID-19 ARDS subjects and 29 historic non-COVID-19 ARDS controls admitted to intensive care units (ICUs) at WCM. There were no significant age, sex or race differences between COVID-19 ARDS (N=31) and non-COVID-19 ARDS subjects (N=29) in the cohort. Cancer was over-represented in the non-COVID-19 ARDS cohort (48.0% versus 3.2% in COVID-19 ARDS). There were also notable differences in respiratory physiology. COVID-19 ARDS was associated with more severe hypoxemia (PaO2:FiO2 ratio, P:F ratio 84 versus 193 in non-COVID-19 ARDS) but lower ventilator ratio (1.65 vs 2.89 in non-COVID-19 ARDS).

The *recovery* cohort included 12 COVID-19 ARDS subjects with plasma available from both their ICU and recovery time point to allow for longitudinal analysis. The median age of this cohort was 47 years old and was majority male (67% versus 33% female).

### Angiopoietin 2 is associated with CD61 staining microthrombi in COVID-19 ARDS subjects

Twenty COVID-19 ARDS lung autopsy specimens were stained for ANGPT2 and CD61 protein. High ANGPT2 protein was associated with increased CD61 (P=0.005, **Supplementary Figure 1**). Representative sections from a high and low ANGPT2 subject are shown in **Figure 2**. ANGPT2 staining was pronounced in the microvasculature and was mirrored by CD61 positive microthrombi in a similar distribution, linking vascular injury and platelet-rich microvascular microthrombi in COVID-19 ARDS.

**Figure 2:**
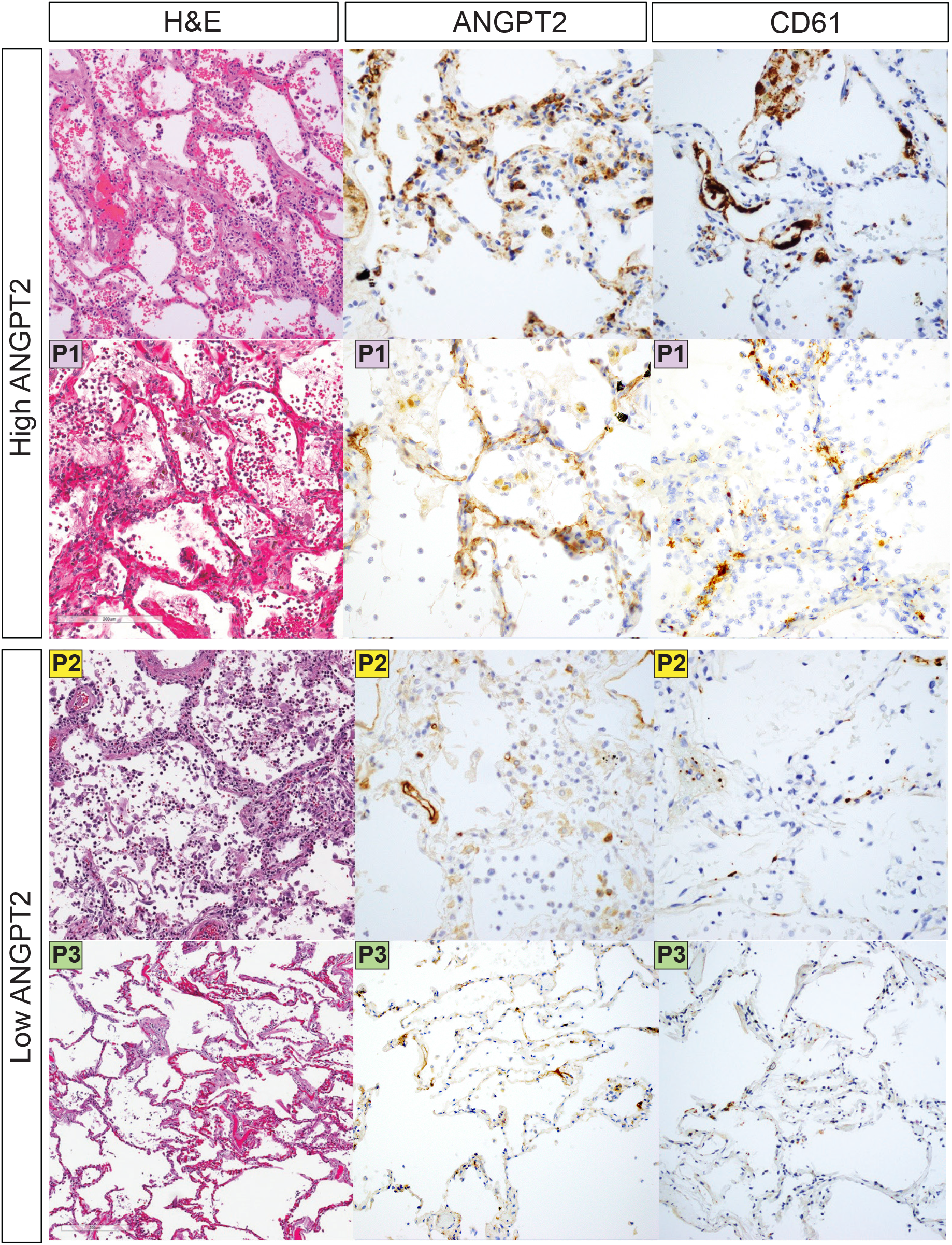
Angiopoietin 2 is associated with CD61 staining microthrombi in COVID-19 ARDS subjects. Angiopoietin-2 (ANGPT2) and CD61 staining in COVID-19 ARDS subjects. Lung autopsy specimens from 20 COVID-19 ARDS subjects were stained for ANGPT2 and CD61. High ANGPT2 (N=10) corresponds to autopsy subjects with ANGPT2 quantification above the median of the autopsy cohort while low ANGPT2 (N=10) represents the low ANGPT2 cohort. High ANGPT2 was associated with increased CD61 staining (P=0.005, Supplementary Figure 1). P1, P2, P3 labels indicate autopsy subjects with serum proteomic data shown in Figure 3A.

### The hospitalized *at risk cohort* blood proteome identifies a maladaptive vascular response preceding critical illness

To test whether the blood proteome would reflect the vascular injury signal seen in the COVID-19 ARDS autopsy specimens, we performed targeted blood proteomics in the *at risk* cohort. Building on the link between vascular injury and platelet-rich microthrombi in the autopsy analysis, we defined a protein set based on the association of circulating proteins with death and platelet levels (**Figure 3A**, see Methods for details on the statistical analysis). We included proteins that significantly associated with both parameters (FDR 0.1): PDGFA, PDGFB, ANGPT1, SORT1, HBEGF, LAP TGFB1, CD84, CXCL5, MMP9, PAI, IL7, IL1RA, and CXCL1. In addition, we selected 9 proteins that were associated with either death or platelet count (FDR 0.1) and have known vascular functions: ADAMTS13, CD40LG, EGFR, SELP, UPA, VEGFA, GP6, and HO1. TIE2 was additionally included since it is the receptor for ANGPT2 (17). The final set comprised 22 proteins (see Methods and **Supplementary Figure 2**), including proteins related to vascular junctional integrity (ANGPT1, TIE2), angiogenesis (PDGFA, PDGFB), platelet degranulation (CD40LG, GP6), and coagulopathy (ADAMTS13, PAI), highlighting the potential functional significance of the identified proteins. Notably, these representative vascular proteins had lower expression in *at risk* subjects who died (**Figure 3B**), representing an early signal of vascular limitation in COVID-19 pathogenesis that we termed the *maladaptive vascular response*.

**Figure 3:**
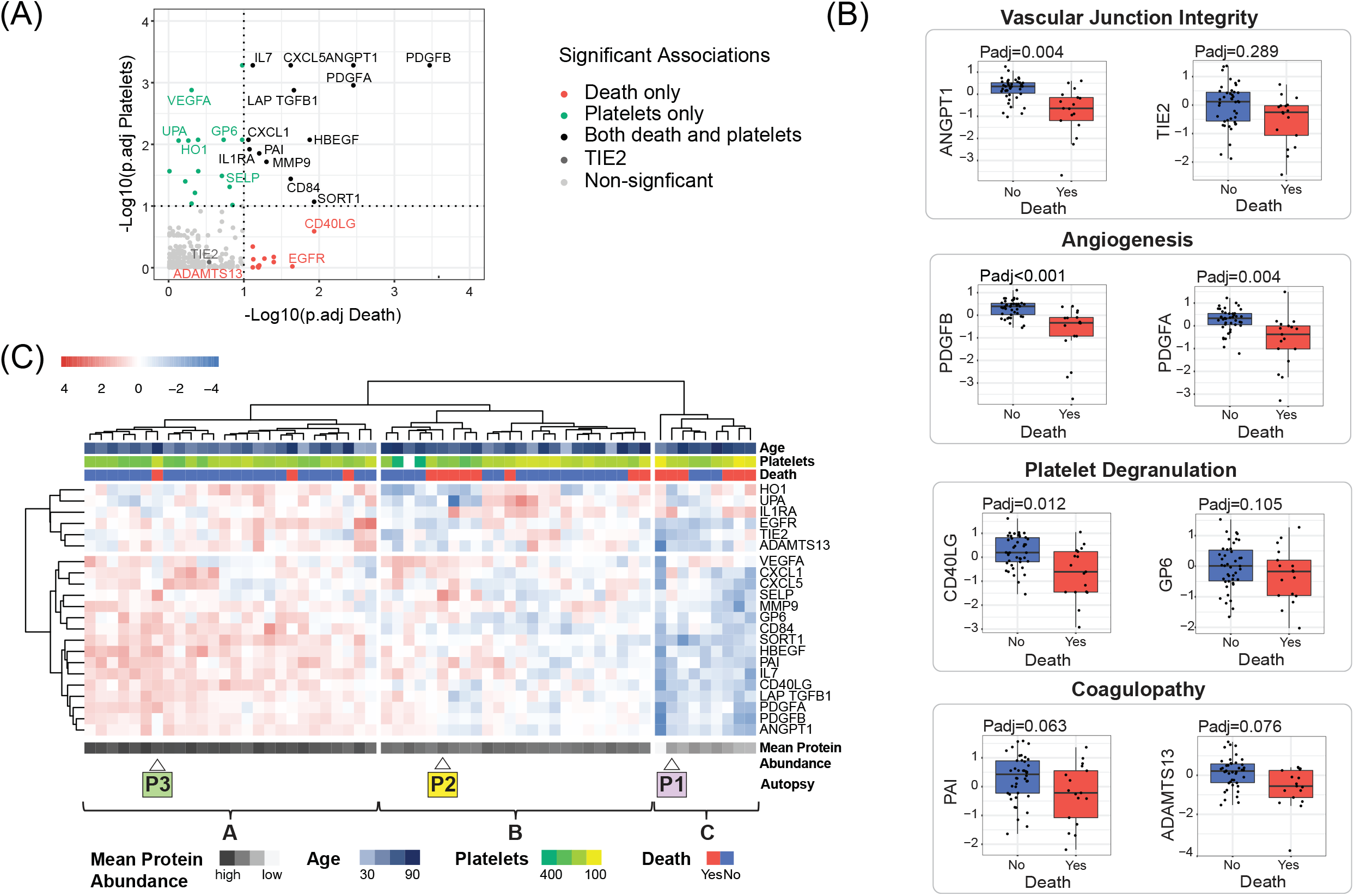
The hospitalized *at risk* cohort (N=59) blood proteome identifies a maladaptive vascular response preceding critical illness. (A) Overview of the assiociations of the protein set to death and platelets. P-values were obtained with a linear mixed effect model to account for repeated measurements and confounders and corrected for multiple comparisons (Padj). X and Y axes indicate the -log10 of the adjusted p-value of the association of proteins to death and platelets, respectively. The protein labels signifies inclusion in the final protein set. (B) Box plots demonstrating the association between proteins of vascular junctional integrity, angiogenesis, platelet degranulation, and coagulopathy to mortality in the *at risk* cohort after adjusting for multiple comparisons. The boxes indicate the interquantile range (IQR) of the data distriibution, the line in the box represents the median value and the whiskers extend for 1.5 times the value of the IQR. Dots indicate the protein level in individual patients. (C) Heatmap of protein set abundance in the *at risk* COVID-19 subjects. Hierarchical clustering was performed using Ward linkage and Euclidean distance. Age, platelet count and death are overlaid at the top. Mean abundance of the 22 protein set and autopsy cases identifiers (e.g. P1, P2, P3) are displayed at the bottom.

Patient clustering based on this protein set identified three distinct patient groups (clusters A, B, and C in **Figure 3C**), with mortality and low platelets progressively enriched. Interestingly, this mortality and low platelet enrichment was associated with lower mean abundance of the 22 proteins (P<0.001, **Supplementary Figure 3A**) and higher age (P=0.016, **Supplementary Figure 3B**). Three of the 20 autopsy subjects were also profiled in this cohort (**Figure 2** and **Figure 3C**, marked **P1, P2, P3**). Autopsy patient P1 (high ANGPT2, high CD61) appears in the lowest protein abundance group (cluster C) while autopsy patient P3 (low ANGPT2, low CD61) can be found in the highest protein abundance group (cluster A), linking ANGPT2 mediated lung vascular injury and CD61 microthrombi to low circulating mean vascular protein abundance.

### Loss of circulating vascular proteins is associated with low platelets, mortality, and plasma ANGPT2 in ARDS

We next tested our set of 22 proteins in the *ARDS* cohort. First, we investigated the vascular protein set in COVID-19 ARDS subjects (**Supplementary Figure 4A**). Confirming the protein results from the COVID-19 *at risk* cohort, low mean protein abundance of the protein set was associated with worse survival (P=0.026, **Supplementary Figure 4B**), low platelet count (P<0.001, **Supplementary Figure 4C**), and older patient age (P=0.035, **Supplementary Figure 4D**). The addition of non-COVID-19 ARDS patients (bacterial sepsis and influenza ARDS), lead to a similar trend (**Figure 4A)** with survival (P=0.020, **Figure 4B**) and low platelets (P<0.001, **Supplementary Figure 5A**) associated with low mean vascular protein abundance (P<0.001, **Supplementary Figure 5B**). Notably, plasma ANGPT2 was higher in the low mean protein abundance cluster (P=0.001, **Figure 4C** and **Supplementary Figure 4E**), linking low vascular protein abundance and plasma ANGPT2 in diverse ARDS subjects.

**Figure 4:**
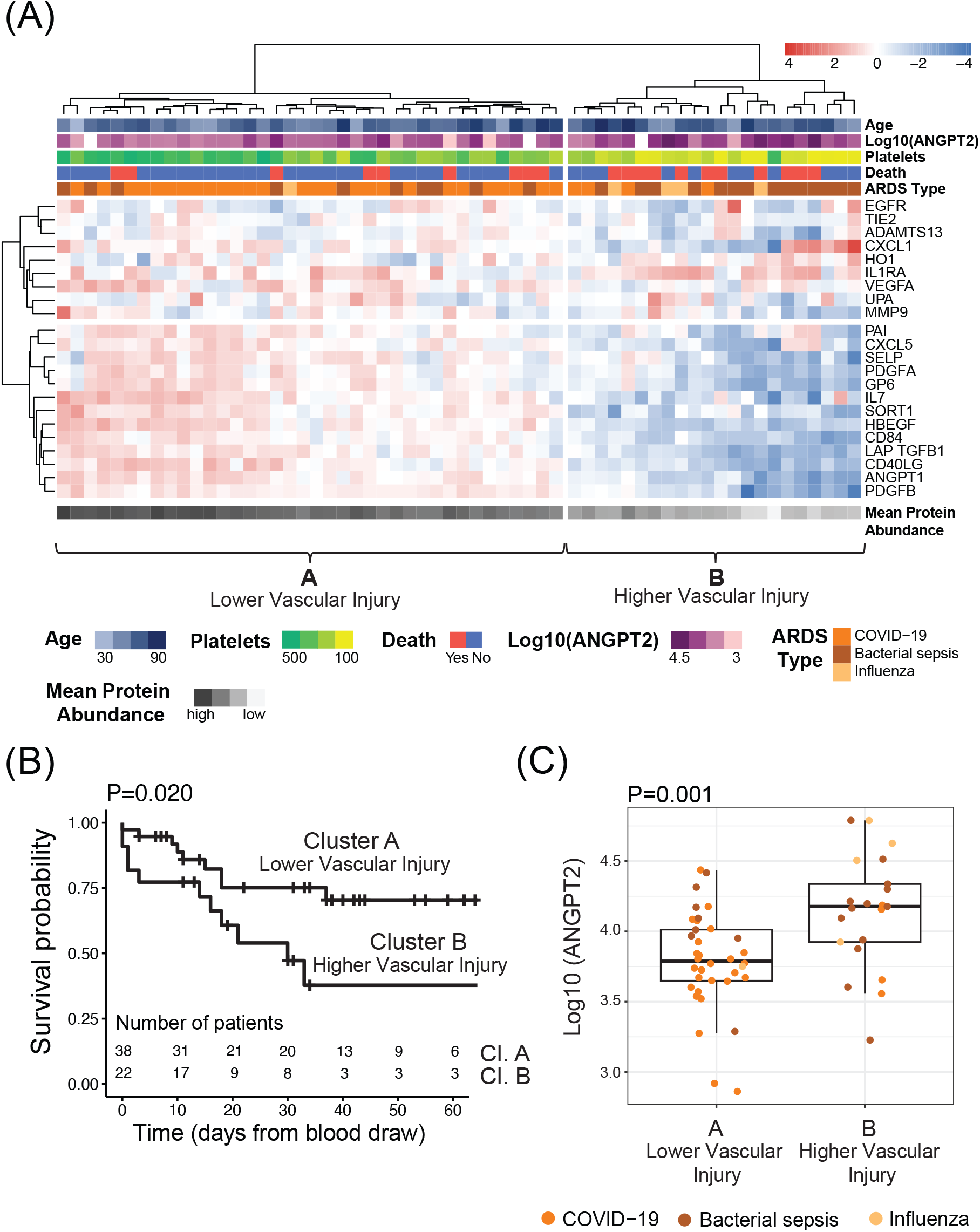
Loss of circulating vascular proteins is associated with thrombocytopenia, mortality, and plasma ANGPT2 in the *ARDS* cohort (N=60). (A) Heatmap of 22 protein set abundance in diverse ARDS subjects, divided into two clusters. Hierarchical clustering was performed using Ward linkage and Euclidean distance. Age, log10(ANGPT2), platelet count, mortality, and ARDS etiology are overlayed at the top. Mean protein abundance of the 22 protein set is overlayed at the bottom. (B) Kaplan-Meier survival analysis for the two heatmap clusters. P-value was estimated using a log-rank test. X-axis was capped at 60 days. The table at the bottom indicates the number of patients at risk at each timepoint in the two clusters. (C) Log10(ANGPT2) values in the two clusters. Differential statistic was assessed with a two-sided Mann-Whitney U test. The boxes indicate the interquantile range (IQR) of the data distribution, the line in the box represents the median value and the whiskers extend for 1.5 times the value of the IQR. Dots indicate the protein level in individual patients across the different ARDS categories: COVID-19 (orange), bacterial sepsis (brown) and influenza (mustard).

Interestingly, when COVID-19 ARDS was considered alone (**Supplementary Figure 4**), this higher vascular injury signature was present in 39% (12 of 31) of COVID-19 ARDS subjects, yet when all three infection types were considered (**Figure 4**), only 13% (4 of 31) of COVID-19 ARDS were in the higher vascular injury cluster compared to 58% (14 of 24) of bacterial sepsis ARDS and 80% (3 of 4) of influenza ARDS subjects, demonstrating that vascular injury may be relative to the causative infection, with COVID-19 ARDS overall being associated with less vascular injury than bacterial sepsis and influenza related ARDS. This finding is supported by a lower ventilator ratio in COVID-19 ARDS subjects compared to non-COVID-19 (**Supplementary Table 1**) a physiologic surrogate for vascular injury in ARDS (18). This is also consistent with previous investigations showing higher platelet counts and less platelet consumption in COVID-19 compared to bacterial sepsis ARDS (19).

### Induction of vascular cell death is associated with ARDS vascular injury

Having validated our vascular injury signature in diverse ARDS populations, we assessed whether ARDS vascular injury could be associated with genetically regulated necrotic cell death, known as necroptosis. We first demonstrated increased expression of plasma RIPK3, a vital necroptosis protein (20), in ARDS subjects with higher vascular injury (P=0.020, **Figure 5A**). Plasma RIPK3 was also correlated with plasma ANGPT2 (r=0.40, P=0.003, **Figure 5B**), supporting the existence of a link between circulating necroptosis proteins and ARDS-related vascular injury. COVID-19 ARDS autopsy subjects demonstrated diffuse microvascular staining for pMLKL, a terminal protein in necrotic cell death execution downstream of RIPK3 (**Figure 5C**), including the high vascular injury autopsy subject P1 (see label **P1** in **Figure 2, Figure 3**, and **Figure 5**), linking induction of necroptosis mediator pMLKL to lung vascular injury and low circulating vascular protein abundance in COVID-19 ARDS.

**Figure 5:**
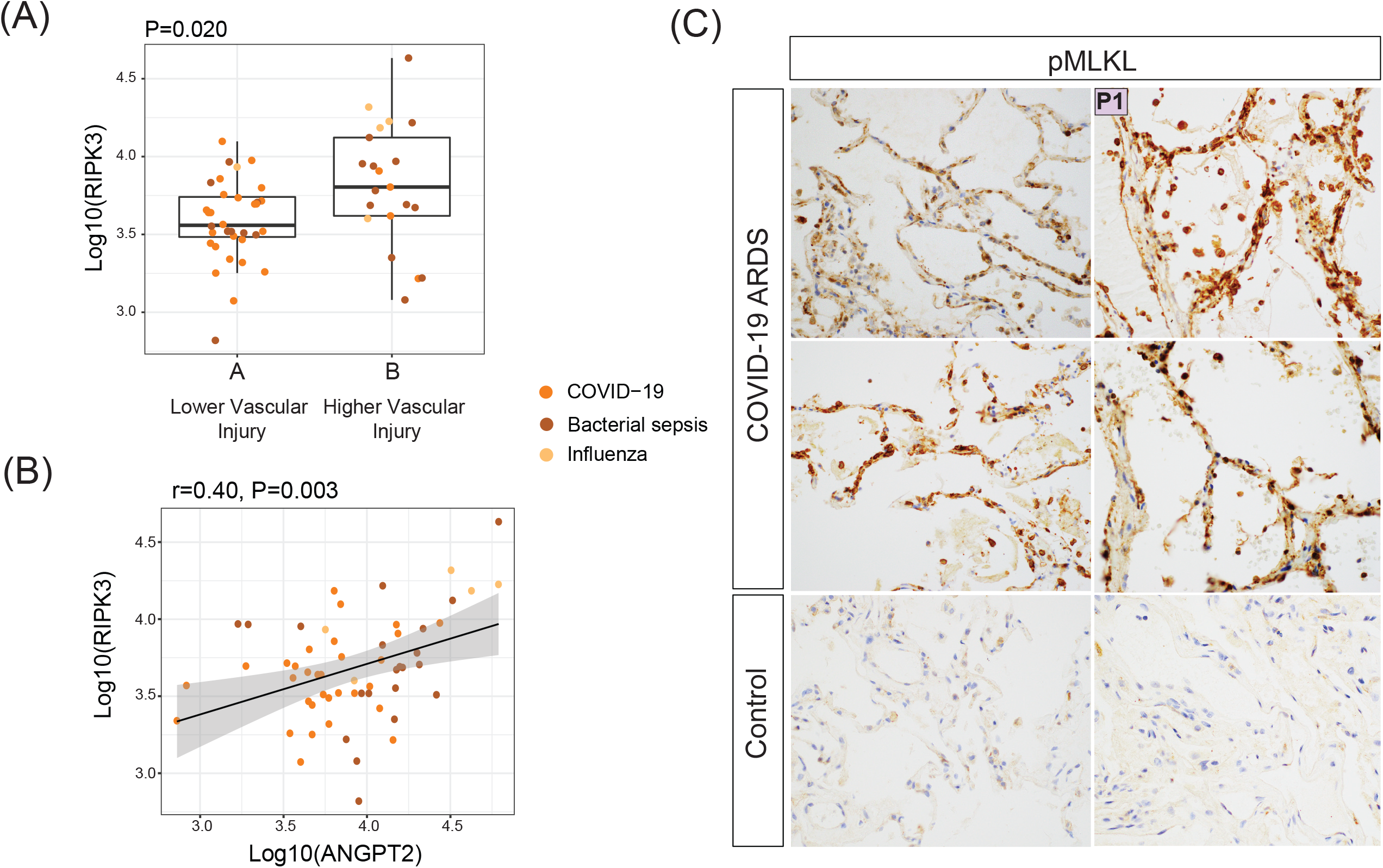
Induction of vascular cell death is associated with ARDS vascular injury. (A) Plasma receptor interacting protein kinase 3 (RIPK3) in ARDS by heatmap cluster (Figure 4A, N=60). Differential statistic was assessed with a two-sided Mann-Whitney U test. The boxes indicate the interquantile range (IQR) of the data distribution, the line in the box represents the median value and the whiskers extend for 1.5 times the value of the IQR. Dots indicate the protein level in individual patients across the different ARDS categories: COVID-19 (orange), bacterial sepsis (brown) and influenza (mustard). (B) Correlation of plasma RIPK3 and plasma ANGPT2 in the ARDS cohort (Figure 4A, N=60). r indicate the Pearson correlation coefficient of the two variable and P its corresponding p-value.The black line represents the linear regression line and the gray area indicates the 95% confidence interval of the fit. Dots indicate the protein level in individual patients across the different ARDS categories: COVID-19 (orange), bacterial sepsis (brown) and influenza (mustard). (C) Phosphorylated mixed lineage kinase domain-like (pMLKL) staining in COVID-19 ARDS autopsy and healthy control subjects. 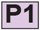 corresponds to autopsy subject from Figure 2 with high ANGPT2 and high CD61 staining and with serum profiling in Figure 3C showing low circulating vascular proteins.

### Among COVID-19 ARDS *recovery* subjects, longitudinal plasma proteomics identifies a stable protein trajectory associated with good functional recovery

We further investigated whether our 22 protein set had predictive ability during recovery. Patient clustering based on the recovery plasma protein set revealed two distinct clusters (**Figure 6A**). Again, the low protein abundance cluster was associated with platelet level (P=0.049, **Supplementary Figure 6A**) and higher age (P=0.049, **Supplementary Figure 6B**). One year follow up functional recovery data based on the EQ-5D-3L questionnaire was available on 11 of these 12 recovery individuals (top annotation in **Figure 6A**, see **Methods** for details). Notably, the cluster of patients with lower abundance of our protein set (P=0.004, **Supplementary Figure 6C**) displayed worse functional recovery 12 months after admission from the ICU, while higher vascular protein abundance was associated with better functional recovery (P=0.027, **Figure 6B**). In order to test whether the protein trajectory from ICU to recovery was different between good and poor functional recovery subjects, we compared the differences in protein abundances between the two timepoints in the two patient clusters (see Methods). For proteins representative of junctional barrier integrity (TIE2, Padj=0.20), angiogenesis (PDGFA, Padj=0.20), platelet degranulation (GP6, Padj=0.20), and coagulopathy (PAI, Padj=0.20), good functional recovery was associated with stable protein trajectory (**Figure 6C**), as opposed to the large protein changes among the poor recovery subjects. This stable trajectory among good functional recovery subjects was similar for platelet levels (P=0.086, **Supplementary Figure 6E**) and ANGPT2 (p=0.083, **Supplementary Figure 6F**).

**Figure 6:**
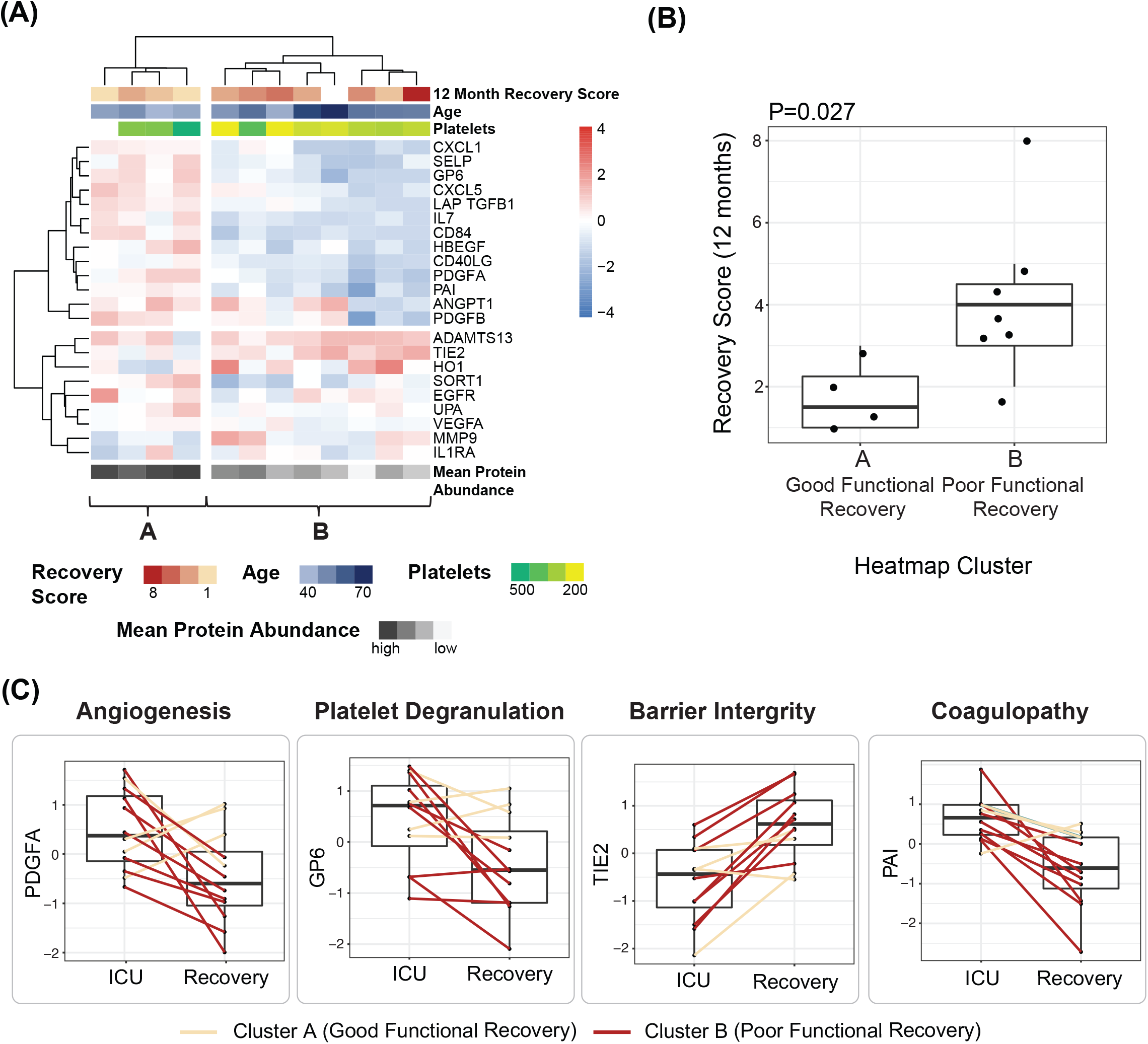
Among COVID-19 ARDS recovery subjects (N=12), longitudinal plasma proteomics identifies a stable protein trajectory associated with good functional recovery. (A) Heatmap of COVID-19 recovery subjects. Functional recovery, age, platelet count and 12 month recovery scores are overlaid at the top. Hierarchical clustering was performed with Ward linkage and Euclidean distance. (B) Follow-up recovery scores at 12 months after ICU admission in the two heatmap clusters. Differential statistic was assessed with a two-sided Mann-Whitney U test. The boxes indicate the interquantile range (IQR) of the data distribution, the line in the box represents the median value and the whiskers extend for 1.5 times the value of the IQR. Dots indicate the protein level in individual patients. High scores indicate worse functional recovery. (C) Trajectory of vascular proteins from ICU to recovery time points by functional recovery group. The boxes indicate the interquantile range (IQR) of the data distribution, the line in the box represents the median value and the whiskers extend for 1.5 times the value of the IQR. Dots indicate the protein level in individual patients in the two timepoints. Values from the same patient are linked by a line and colored according to the corresponding heatmap cluster: A (cream) or B (red). Differential statistic of the protein trajectories between the two patient clusters was computed with a linear model. All displayed trajectory differences were significant to an adjusted p-value<0.25.

## DISCUSSION

In this study, we traced a maladaptive vascular response through the natural history of COVID-19 ARDS from hospital admission to either recovery or death. Reflected in both the lung tissue and blood proteome, we demonstrated the clinical relevance of the low abundance of circulating vascular proteins with known vascular functions and implied a link with vascular cell death, and in particular specialized necroptotic cell death.

This vascular phenotype is notably present in certain COVID-19 subjects prior to ICU admission. While vascular injury spans the COVID-19 disease continuum from asymptomatic blue toes to catastrophic thromboembolic disease and ARDS-associated microangiopathy, our identification of broad loss of vascular signaling in early severe disease generalizes this maladaptive vascular response to the large population of hospitalized COVID-19 subjects. The loss of vascular proteins could result from SARS-CoV-2 endothelial infection (1, 2), although this remains controversial and thus far only reproducible in artificially engineered endothelial cell (21), while primary human endothelial cell appear resistant to infection (22). Alternatively, in common with bacterial sepsis (16, 23, 24) and influenza infection (25), unrestrained COVID-19 related inflammatory signaling (9) could similarly induce vascular cell death. Indeed, we demonstrate induction of genetically regulated necrotic cell death mediator (pMLKL) in the microvasculature of high vascular injury COVID-19 autopsy subjects. Diverse upstream mediators previously linked to COVID-19 (e.g. TNF-alpha (26), interferons (27–31)) can induce necroptosis (20), providing a crucial link between SARS-CoV-2 infection and both direct (virus) or indirect (TNF, interferons) induction of vascular cell death in COVID-19 subjects.

The role of activated platelets in vascular injury and repair is also apparent in our data. Activated platelets amplify immune responses in early ARDS but also play an essential role in vascular repair. The consistently low platelet levels across our cohorts and the extensive microthrombi observed in our autopsy subjects implies a circulating milieu of platelet consumption. This milieu of platelet consumption is supported by a blood signature of ongoing thrombolysis (high UPA and low PAI) and low levels of platelet derived proteins (low SELP, and GP6) in our high vascular injury subjects. Relative loss of ADAMTS13, linked to secondary microangiopathy in COVID-19 (32), is similarly deficient in our higher vascular injury subjects, linking platelet consumption with microangiopathy in severe COVID-19. Low platelets have previously been linked to ARDS mortality (33) and our data suggest this may be related to depletion in platelet related angiogenic (34–36) and junctional barrier factors (37–40). Consistently low circulating angiogenic (low PDGFA and PDGFB) and barrier protein (low ANGPT1) in our higher vascular injury and low platelet subjects imply limitations in these essential reparative processes.

The validation of our vascular phenotype across diverse causes of ARDS broadens the relevance of our findings. In linking low platelets, vascular function, and mortality in COVID-19, bacterial sepsis, and influenza ARDS, we hint at a common final pathway of vascular injury that is more disease- (ARDS) than cause- (COVID-19) specific. Of note is that this vascular injury pattern may be related to a reduced baseline vascular resilience in our high vascular injury subjects. Consistently, our high vascular injury subjects are older (41), have worse baseline renal function (42, 43), and are more likely to have cancer (44) (**Supplementary Table 2**), all variables know to be associated with vascular disease.

The identification of this severe vascular phenotype across infectious causes of ARDS also presents an opportunity for targeted vascular therapies in ARDS, including those that have shown promise in COVID-19 (45), ARDS generally (46), and in exciting preclinical (47, 48) and early human experimental therapies, including ANGPT1 supplementation trial currently underway in COVID-19 subjects (49). And while a ANGPT2 neutralizing antibody study in hospitalized patient with COVID-19 was stopped for futility in October 2020 (50), our data could improve patient selection for similar trials in the future, including the use of platelet levels to identify subjects with vascular limitation.

Finally, our identification of a vascular recovery proteome is novel. An estimated 2 million patients have been hospitalized in the United States since the start of the COVID-19 pandemic, with the overwhelming majority recovering (51). But even in recovery, patients remain at risk for disease related morbidity and mortality (52). We demonstrate that a stable circulating vascular proteome is important for functional recovery. This association between vascular stability, platelet levels, and functional recovery could also support platelet levels as a novel biomarker in ARDS recovery. Larger studies will be needed to validate this observation.

In summary, we identify an early vascular injury signal in COVID-19 ARDS that has predictive value in early disease through to recovery and well as in bacterial sepsis and influenza ARDS and could improve patient selection and timing of vascular targeted therapies in ARDS.

## METHODS

### Study design

This study enrolled COVID-19 subjects at New York Presbyterian Weill Cornell Medical Center (WCM) between March 15 and August 17, 2020 with blood specimens obtained during routine care and as part of existing study protocols. Additional historic non-COVID-19 ARDS samples from influenza and bacterial ARDS patients prospectively enrolled into the Weill Cornell Biobank of Critical Illness (BOCI) from October 20, 2014 until May 24, 2020 were included as part of the *ARDS* cohort. COVID-19 study samples were analyzed according to ARDS status (*at risk, ARDS* or *recovery*) at study enrollment. The *at risk* cohort included 59 adult (>18) non-pregnant COVID-19 subjects admitted to the general wards of WCM with serum available and who did not meet ARDS criteria at study enrollment. The *ARDS* cohort included adult (>18) non-pregnant COVID-19 (N=31) and historic non-COVID ARDS (N=29) subjects admitted to the intensive care unit (ICU) at WCM. For the *ARDS* cohort, only study subjects meeting ARDS criteria and with blood sampling within 10 days of ICU admission were considered for analysis. The *recovery* cohort included 12 adult (>18) non-pregnant COVID-19 ARDS subjects with plasma samples available from both the time of ICU care and the subsequent recovery period to allow for longitudinal analyses. *Recovery* blood samples were obtained from patients convalescing in the hospital rehabilitation floors, as well as from the New York Presbyterian Weill Cornell Medicine Post-ICU recovery clinic.

### Study Approval

The study was approved by the institutional review board (IRB) at WCM (20-05022072, 20-0302168, 20-03021681, and 1811019771). Written informed consent was received from all participants prior to inclusion in the study.

### Blood sampling

In the *at risk* cohort, between 1 and 3 consecutive daily samples were obtained from our central lab after routine processing to obtain serum. To obtain serum, blood collected in serum separator tubes (SST) was processed within 2 hours of venipuncture. Whole blood was centrifuged at 1,500 g for 7 minutes. The serum layer was aliquoted and stored at -80°C. These samples were obtained with a waiver of informed consent. In this cohort, samples collected after patient intubation were excluded from the analysis. In the *ARDS* and *recovery* cohorts, plasma was isolated from study subjects according to our existing plasma isolation protocol (53–56). To obtain plasma, blood collected in EDTA tubes was processed within 6 hours of venipuncture. Whole blood was centrifuged at 490 g for 10 minutes. The plasma layer was removed in 200 uL aliquots and stored at -80.

### Clinical evaluation

Baseline clinical parameters and outcomes were extracted from the electronic medical record (EMR) as described previously (57, 58). Baseline comorbidities were manually abstracted from the EMR. Baseline clinical data (labs, severity of illness, ventilator data) were measured at time of blood sampling in both the *at risk* cohort and *ARDS* cohort. Severity of illness was defined by the sequential organ failure assessment score (SOFA) (59). ARDS was determined according to the Berlin definition with ARDS severity capped at mild for subjects on non-invasive ventilation (60). Two critical care investigators independently adjudicated the ARDS diagnosis. In all study subjects, COVID-19 was diagnosed if a subject had a syndrome compatible with COVID-19 and a nasopharangeal (NP) swab positive for SARS-CoV-2 by reverse transcriptase polymerase chain reaction (RT-PCR).

### Recovery Evaluation

*Recovery* subjects were assessed for recovery using the EuroQol-5D-3L (EQ-5D-3L) questionnaire (61) at 12 months after ICU admission. The EQ-5D-3L is a self-assessment of the patient recovery, and considers 5 distinct domains, namely mobility, self-care, usual activities, pain or discomfort, and anxiety or depression (62). Each domain was scored 0, 1, or 2 depending on whether the patient reported no, some, or extensive limitations in each respective domain. For each patient, a final score was defined as the sum of the scores across the five domains and treated as an ordinal variable in the statistical analysis. Maximal functional limitation would have a score of (2*5=)10 while an optimal recovery would be scored 0.

### Autopsy studies

Twenty autopsies performed between March 19 and June 30, 2020 with pre-mortem nasopharyngeal swabs positive for SARS-CoV-2 were considered for lung tissue staining. Lung tissue specimens were fixed in 10% formalin for 48–72 hours. Hematoxylin and eosin staining were performed for all cases. Immunohistochemistry was carried out for angiopoietin-2 (sc-74403, Santa Cruz, TX, 1:100), CD-61 (CD61 clone 2F2, Leica Biosystems, IL) and phosphorylated mixed lineage kinase domain-like (pMLKL, MAB91871, NOVUS Biologicals, CO, 1:750 with casein for background reduction). Specimens were scanned by whole-slide image technique using an Aperio slide scanner with a resolution of 0.24 μm/pixel. Control tissue was from non-diseased sections of lung taken during clinically indicated lung biopsies. Quantification of ANGPT2 and CD61 was performed on four random 20X images selected using a random overlay of points and excluding fields with large vessels or airway. All twenty autopsies were analyzed using Immunohistochemistry profiler (63) as a plugin for Image J (National Institutes of Health, USA). High, intermediate, low, and overall percent positive was averaged over the four measurements. The median ANGPT2 quantification was used to define the high (>median) and low (<median) ANGPT2 staining. The association between CD61 and ANGPT2 was then calculated based on CD61 quantification in the low and high ANGPT2 groups using Mann-Whitney U test for continuous variables.

### O-link Plasma Proteomics

Plasma and serum samples from the *at risk, ARDS* and *recovery* cohorts were profiled using O-Link through the Proteomics Core of Weill Cornell Medicine-Qatar. The O-link assays were performed using Inflammation (v.3021), Cardiovascular II (v.5005), and Cardiovascular III (v.6113) panels (O-link, Uppsala, Sweden). EDTA plasma and serum samples were heat-inactivated at 56 degrees for 15 mins according to the virus inactivation protocol provided by O-link) (64). The protein measurements were performed with the Proximity Extension Assay technology (PEA) according to manufacturer’s instructions. In summary, high throughput real-time PCR of reporter DNA linked to protein specific antibodies was performed on a 96-well integrated fluidic circuits chip (Fluidigm, San Francisco, CA). Signal quantification was carried out on a Biomark HD system (Fluidigm, San Francisco, CA). Each sample was spiked with quality controls to monitor the incubation, extension, and detection steps of the assay. Additionally, samples representing external, negative and inter-plate controls were included in each analysis run. From raw data, real time PCR cycle threshold (Ct) values were extracted using the Fluidigm RT-PCR analysis software at a quality threshold of 0.5 and linear baseline correction. Ct values were further processed using the O-link NPX manager software (O-link, Uppsala, Sweden). Here, log2-transformed Ct values from each sample and analyte were normalized based on spiked-in extension controls and scale-inverted to obtain normalized log2 scaled Protein eXpression (NPX) values. NPX values were further adjusted based on the median of inter plate controls (IPC) for each protein and intensity median scaled between all samples and plates.

The *at risk* cohort was profiled in two separate runs. The second run included a total of 11 samples, among which 5 bridge samples were used to scale this batch toward the first one, as recommended by Olink. First, for each bridge sample, the pairwise difference between the first and second batch was computed. An overall batch adjustment factor was then derived as the median of these pairwise differences and subtracted to the values in the second batch.

Subsequently, protein levels were exponentiated, normalized using probabilistic quotient normalization (65) and log2-retransformed. Missing values were imputed using a k-nearest neighbors approach (66) (k=10). 10 proteins were measured across multiple panels and, therefore, their duplicated values were averaged, leaving a total of 266 unique proteins. Protein values were standardized prior to statistical analysis.

### Protein subset derivation

The protein vascular signature was derived in the *at risk* cohort. First, we associated each of the 266 measured proteins to death and platelet count, respectively. Then, we selected proteins associated with both outcomes (adjusted p-value=<0.1, see Statistical Analysis section for details). Additionally, we included proteins associated with either mortality or platelet count (adjusted p-value=<0.1) and with known, well characterized links to vascular function. TIE2 was additionally included as it is the receptor for ANGPT2(17).

### ELISA measurements

Plasma samples from the *ARDS* and *Recovery* cohorts were used for enzyme-linked immunosorbent assays (ELISA) according to manufacturer recommendations. Human ANGPT2 (R&D, CAT#DANG20) and receptor interacting protein kinase 3 (RIPK3, Cusabio, CAT#CSB-EL019737HU) kits were used to measure plasma protein levels. Plasma samples were diluted (1:8 dilution for ANGPT2, 1:10 for RIPK3) prior to plating. Final sample absorbance was measured at 450 nm with wavelength correction performed at 570 nm. Sample concentrations were calculated from a four-parameter logistic curve created from known standard concentrations. Dilution factors were accounted for to calculate the final sample concentration. Plasma ANGPT2 and RIPK3 values were log10-transformed prior to statistical analysis.

### Statistics

In the *at risk* cohort proteomic analysis, protein associations to death (i.e. whether the patient ended up dying) and platelet count (minimum value across the sampling days) were computed using a mixed linear effect model, which allows to properly account for the multiple samples collected per patient. The model was formulated as follows: *protein ∼ outcome + replicate + batch + (1*|*patient)*, where *outcome* was either death or platelet count, *replicate* indicated the day of blood sample draw (first, second or third since hospital admission), and *batch* indicated whether the sample was measured in the first or second run. Association p-values were corrected for multiple testing using the Benjamini-Hochberg method for controlling the false discovery rate (67). Adjusted p-values less than 0.1 were considered significant.

For all cohorts, patient hierarchical clustering based on the standardized proteomics value was performed using Ward linkage and Euclidean distance. The differential analysis between patient clusters was performed using Mann-Whitney U tests for continuous variables, Kendall’s rank correlation for ordinal variables, and log-rank tests for survival times. The correlation between ANGPT2 and RIPK3 was estimated using Pearson correlation. For these analyses, a p-value of less than 0.05 was considered significant.

In the recovery cohort, we first divided patients into two groups based on unsupervised hierarchical clustering (Ward linkage, Euclidean distance) performed on the recovery timepoint. Then, for each patient we calculated the protein abundance difference (*delta*) between the ICU and recovery timepoints. Finally, for each protein we investigated whether the protein delta was different across the two patient groups using the linear model *delta ∼ group*. P-values were corrected for multiple tests using the Benjamini-Hochberg method. Given the small sample size and validation of protein set in two prior cohort, we considered an adjusted p-value less than 0.25 as significant.

All statistical analyses were performed in R 4.0.1. The R code used to generate the statistical findings presented in this paper is publicly available at https://github.com/krumsieklab/covid-vascular-injury.

## Data Availability

The datasets used for this study include sensitive patient information extracted from the electronic health record. They are therefore subject to federal legislation that limits our ability to make them publicly available, even after being subjected to deidentification techniques. To request access to the de-identified minimal datasets underlying the findings illustrated in our paper, interested and qualified researchers should contact Information Technologies & Services Department of Weill Cornell Medicine.
The R Code used to generate all the statistical results presented in this paper is available at https://github.com/krumsieklab/covid-vascular-injury.

https://github.com/krumsieklab/covid-vascular-injury

## Data and Code Availability

The datasets used for this study include sensitive patient information extracted from the electronic health record. They are therefore subject to federal legislation that limits our ability to make them publicly available, even after being subjected to deidentification techniques. To request access to the de-identified minimal datasets underlying the findings illustrated in our paper, interested and qualified researchers should contact Information Technologies & Services Department of Weill Cornell Medicine.

The R Code used to generate all the statistical results presented in this paper is available at https://github.com/krumsieklab/covid-vascular-injury.

## AUTHOR CONTRIBUTIONS

DRP, EB share the first author position. DRP is listed first based on higher total effort to the project. DRP, EB, JK, AMKC designed the study. DRP, ACR, and ACB performed the autopsy staining analyses. LGE, SAM, AC, CNP, AR, JGC, SZJ processed samples and organized the patient clinical data. EB, HS, RB, MB, KC, FS, JK analyzed the proteomic data. KLH and IE provided statistical support for patient clinical data. EL, KW, CNP, LL perform functional assessment of recovery subjects. DRP, EB, JK, AMKC, RB, FS, JGC, EJS, ACR, HOR, JCL, MEC, and SR critically appraised the final dataset. DRP, EB wrote the manuscript. All authors approved the final manuscript.

## ACKNOWLEDGEMENTS

This work is supported by the Biomedical Research Program at Weill Cornell Medicine in Qatar, a program funded by the Qatar Foundation. JK is supported by the National Institute of Aging of the National Institutes of Health under award 1U19AG063744. The authors thank Ilias Siempos for critically reviewing the manuscript.

