## Supplementary Tables and Figures for "The maladaptive vascular response in COVID-19 acute respiratory distress syndrome and recovery"

Supplementary Table 1: Baseline Characteristics and Intensive Care Unit Outcomes of Cohorts

| | At Risk Cohort[A] | ARDS Cohort[A] | | Recovery Cohort[A,B] |
|---|---|---|---|---|
| **Characteristics** | COVID-19 | COVID-19 ARDS | Non-COVID-19 ARDS | COVID-19 |
| **N** | 59 | 31 | 29 | 12 |
| **Age** | 69 (59, 78) | 62 (50, 70) | 60 (52, 70) | 47 (39, 52) |
| **Sex** | | | | |
| Female | 21 (36%) | 6 (19%) | 12 (41%) | 4 (33%) |
| Male | 38 (64%) | 25 (81%) | 17 (59%) | 8 (67%) |
| **Race** | | | | |
| Asian | 2 (3.4%) | 4 (13%) | 3 (10%) | 0 (0%) |
| Black | 9 (15%) | 4 (13%) | 2 (6.9%) | 1 (8.3%) |
| Other or unknown | 21 (36%) | 13 (42%) | 14 (48%) | 9 (75%) |
| White | 27 (46%) | 10 (32%) | 10 (34%) | 2 (17%) |
| **Co-morbidities** | | | | |
| BMI | 28 (23, 31) | 27 (25, 33) | 26 (21, 28) | 28 (26, 30) |
| Hypertension | 31 (53%) | 14 (45%) | 12 (41%) | 4 (33%) |
| CAD | 11 (19%) | 3 (9.7%) | 4 (14%) | 0 (0%) |
| CVA | 7 (12%) | 2 (6.5%) | 0 (0%) | 0 (0%) |
| Heart Failure | 7 (12%) | 1 (3.2%) | 2 (6.9%) | 0 (0%) |
| CKD | 3 (5.1%) | 0 (0%) | 4 (14%) | 0 (0%) |
| Cancer | 9 (15%) | 1 (3.2%) | 14 (48%) | 0 (0%) |
| **Severity of Illness** | | | | |
| SOFA | | 10 (8, 12) | 10 (9, 12) | 9 (8, 10) |
| **Labs** | | | | |
| White blood cell | 5 (4, 7) | 8 (8, 12) | 12 (5, 21) | 9 (7, 10) |
| Lymphocyte Count | 0.7 (0.4, 0.9) | 0.7 (0.5, 1.3) | Missing | 0.8 (0.6, 1.2) |
| Platelet count | 152 (123, 217) | 319 (219, 390) | 122 (54,246) | 312 (236, 411) |
| D Dimer | 389 (220, 653) | 884 (580, 2,100) | Missing | 1,576 (739, 2,515) |
| Fibrinogen | 515 (461, 582) | 749 (562, 919) | 566 (452, 672) | 699 (506, 743) |
| aPTT | 32 (29, 34) | 33 (30, 41) | 33 (28,44) | 30 (27, 32) |
| PT | 13 (13, 15) | 15 (13, 17) | Missing | 13 (12, 14) |
| Creatinine | 0.8 (0.7, 1.1) | 0.9 (0.6, 1.4) | 2.0 (1.3, 3.4) | 0.6 (0.5, 0.9) |
| **Ventilator Parameters** | | | | |
| FiO2 | | 70 (60, 90) | 100 (70, 100) | 50 (40, 70) |
| P:F ratio | | 84 (68, 112) | 193 (132, 272) | 98 (78, 136) |
| Ventilator ratio | | 1.65 (1.46, 2.07) | 2.89 (2.41, 3.44) | 1.90 (1.34, 1.95) |
| Plateau pressure | | 27.0 (24.0, 29.0) | 25.0 (20.0, 28.0) | 26.5 (21.5, 28.5) |
| PEEP | | 14.0 (10.0, 14.5) | 7.2 (5.0, 10.0) | 11.0 (10.0, 13.5) |
| Tidal volume exhaled (ml) | | 431 (382, 472) | 664 (545, 841) | 430 (366, 452) |
| Exhaled minute ventilation | | 10.9 (8.8, 12.5) | 15.2 (12.6, 15.9) | 10.4 (8.8, 11.0) |
| Driving pressure | | 12.5 (12.0, 16.5) | 14.5 (12.0, 17.8) | 13.5 (10.8, 16.2) |
| **Intensive Care Unit 60 Day Outcomes** | | | | |
| Ventilator-Free Days | | 4 (0, 14) | 7 (0, 23) | 6 (0, 14) |
| Mortality | | 8 (26%) | 12 (41%) | 0 (0%) |

[A] Median (IQR). [B] *Recovery* variables are from intensive care unit timepoint to allow direct comparison to the *ARDS* cohort. BMI = body mass index, CAD = coronary artery disease, CVA = cerebral vascular event, CKD = chronic kidney disease, SOFA = sequential organ failure assessment score, FiO2 = fraction of inspired oxygen, P:F ratio = partial pressure of arterial oxygen to fraction of inspired oxygen, PEEP = positive end expiratory pressure.

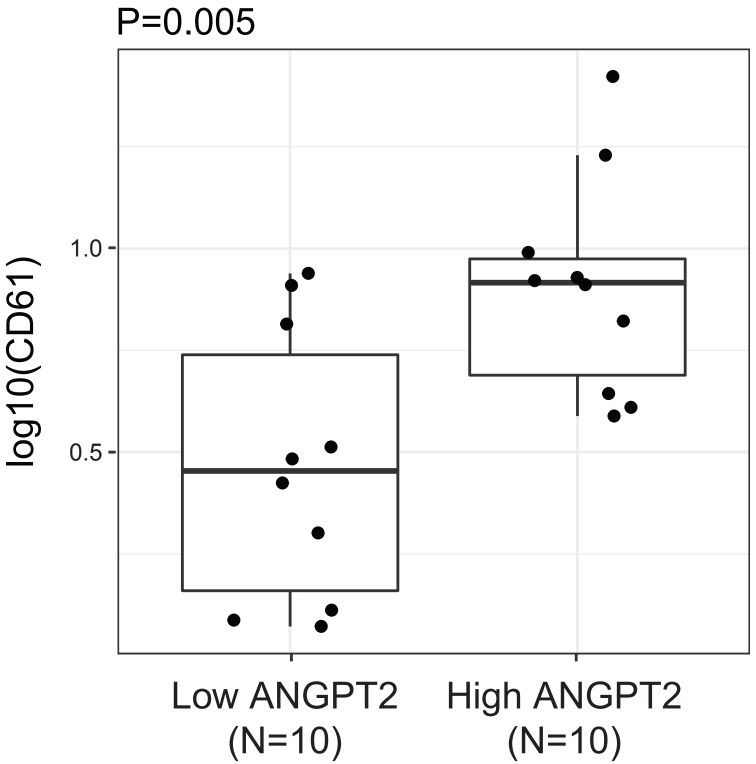


**Supplementary Figure 1: ANGPT2 and CD61 quanfication in COVID-19 autopsy cohort (N=20).** Quantification was performed using Immunohisto-chemistry profiler as a plugin for ImageJ. High and low ANGPT2 corresponds to the autopsy subjects with ANGPT2 staining above and below the median quanti-fication, respectively. The boxes indicate the interquantile range (IQR) of the data distribution, the line in the box represents the median value and the whis-kers extend for 1.5 times the value of the IQR. Dots indicate the protein level in individual patients. Differential statistic was assessed with a two-sided Mann-Whitney U test.

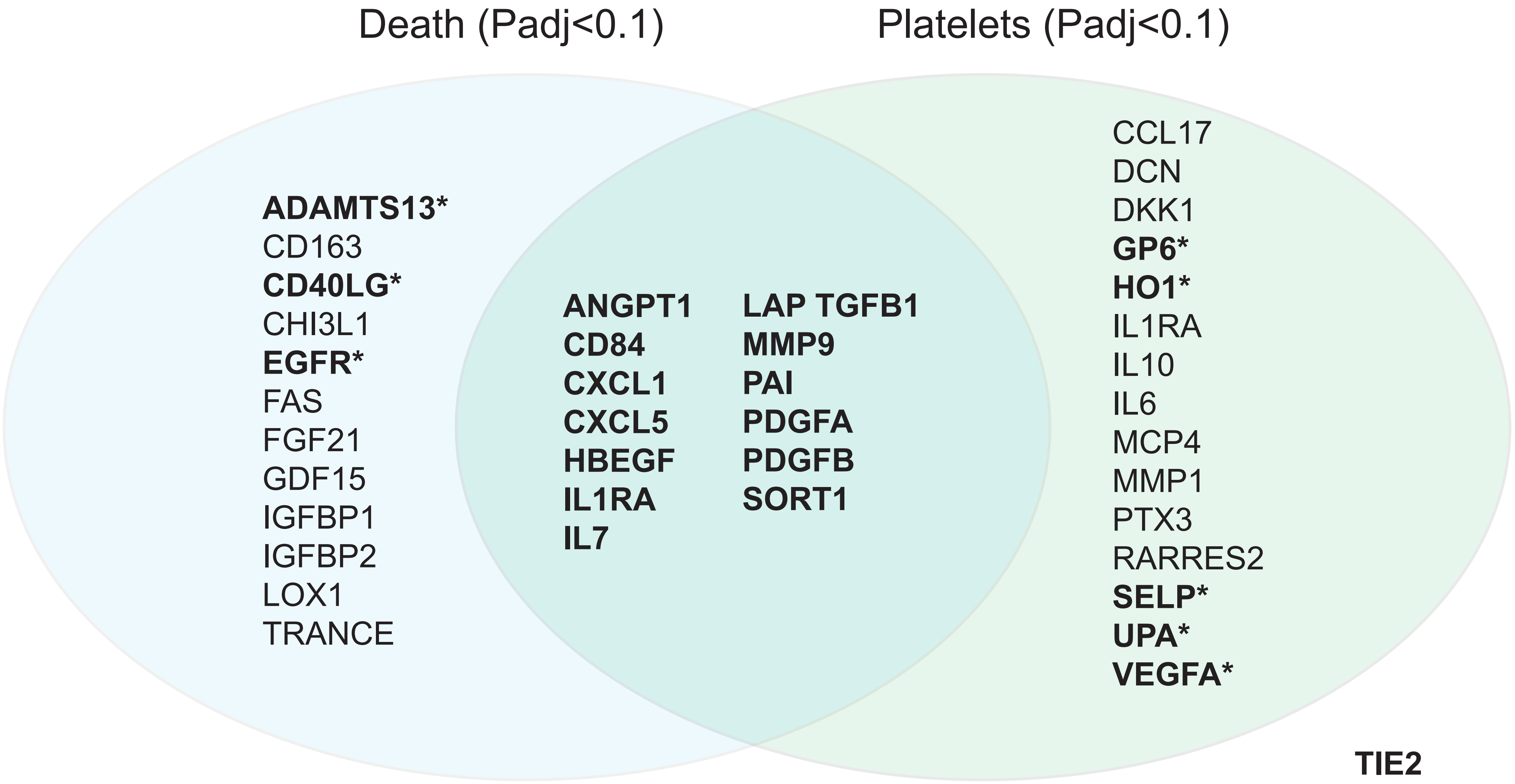


**Supplementary Figure 2: Protein subset selection.** Proteins in bold represent the final protein subset. To identify these proteins, we first considered proteins associated with both death (Padj<0.1) and platelet level (Padj<0.1), indicated in the overlap region. Additionally, proteins associated with either death or platelets with know essential vascular functions were included (ADAMTS13, CD40LG, EGFR, SELP, UPA, VEGFA). These proteins are annotated with an asterisk. Finally, TIE2 was included in the final subset based on it being the receptor for ANGPT2.

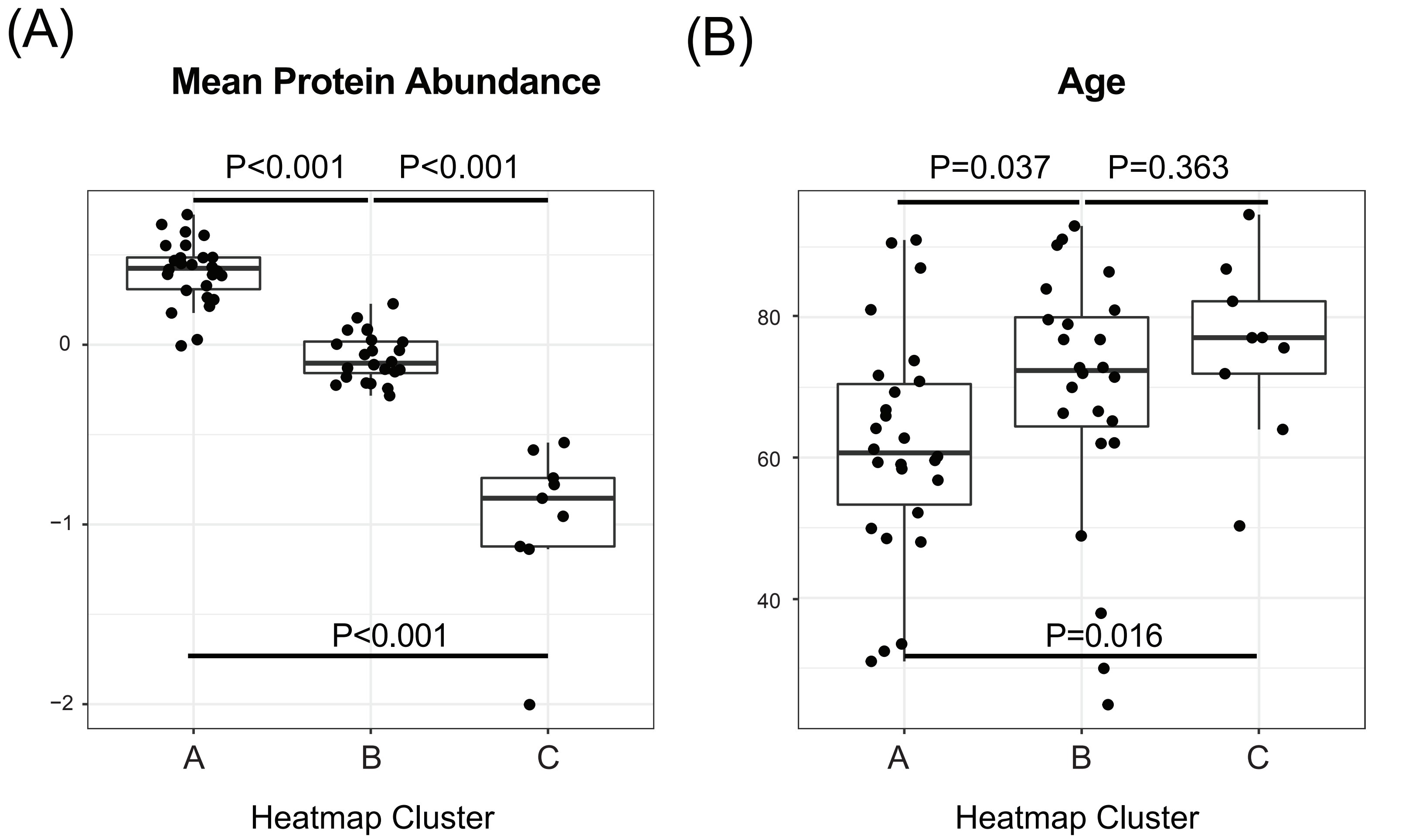


**Supplementary Figure 3: Mean protein abundance and age by heatmap cluster in the *at risk* cohort (N=59).** (A) Successive protein clusters demonstrate significant loss of protein set expression (P<0.001). (B) At risk protein clusters identify significant age differences (P=0.016). The boxes indicate the interquantile range (IQR) of the data distribution, the line in the box represents the median value and the whiskers extend for 1.5 times the value of the IQR. Dots indicate the protein level in individual patients. Differential statistic was assessed with a two-sided Mann-Whitney U test.

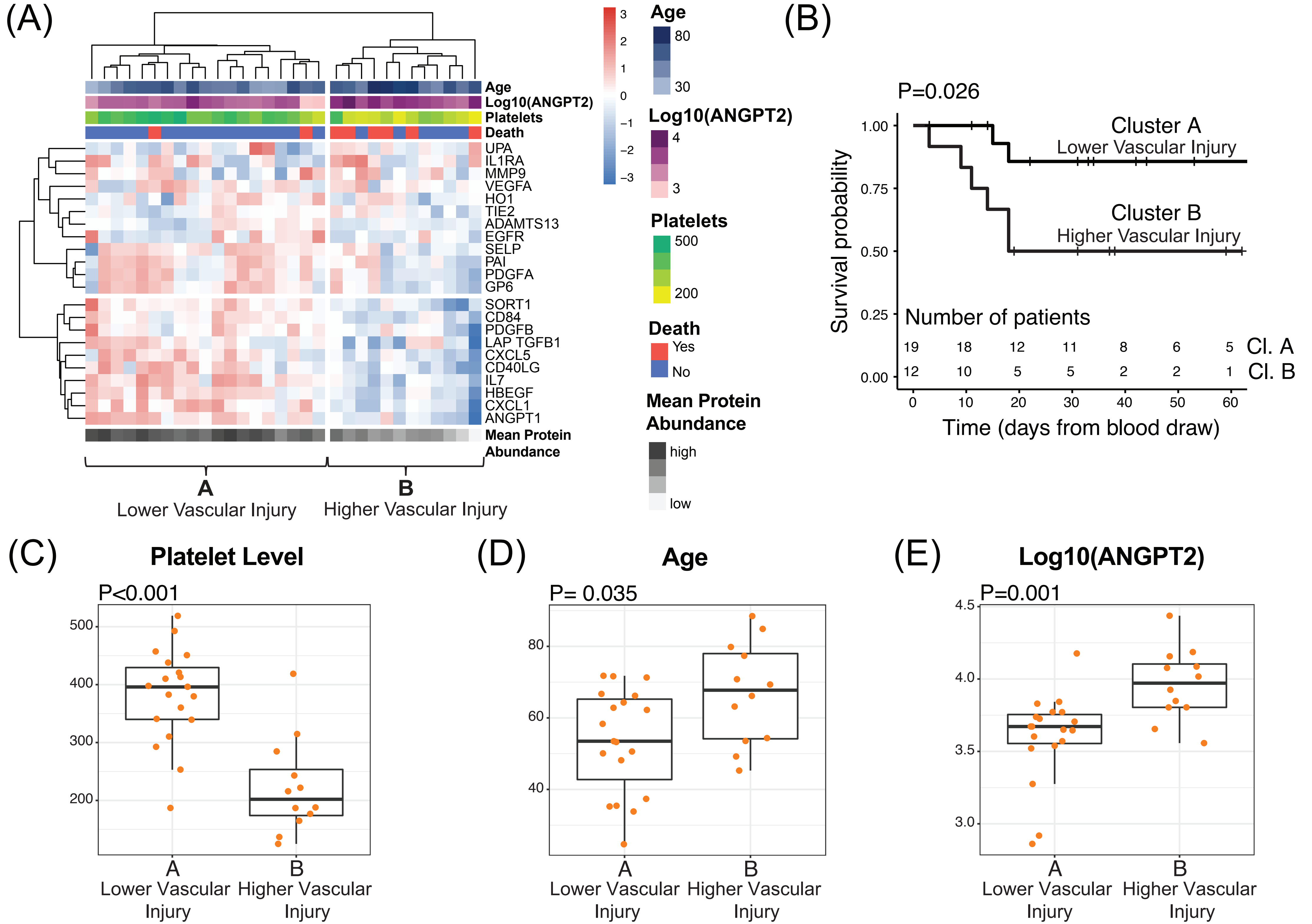


**Supplementary Figure 4: Loss of essential vascular proteins is associated with thrombocytopenia, mortality, and ANGPT2 in COVID-19 ARDS (N=31).** (A) Heatmap of protein subset in COVID-19 ARDS subjects. Age, log10(ANGPT2), platelet count, and mortality are overlayed at the top. (B) Kaplan-Meier plot for the two heatmap clusters. P-value was estimated with a log-rank test. X-axis was capped at 60 days. The table at the bottom indicates the number of patients at risk at each timepoint in the two clusters. (C-E) Platelet levels, age, and Log10(ANGPT2) values in the two heatmap clusters, respectively. The boxes indicate the interquantile range (IQR) of the data distribution, the line in the box represents the median value and the whiskers extend for 1.5 times the value of the IQR. Dots indicate the protein level in individual patients. Differential statistic was assessed with a two-sided Mann-Whitney U test.

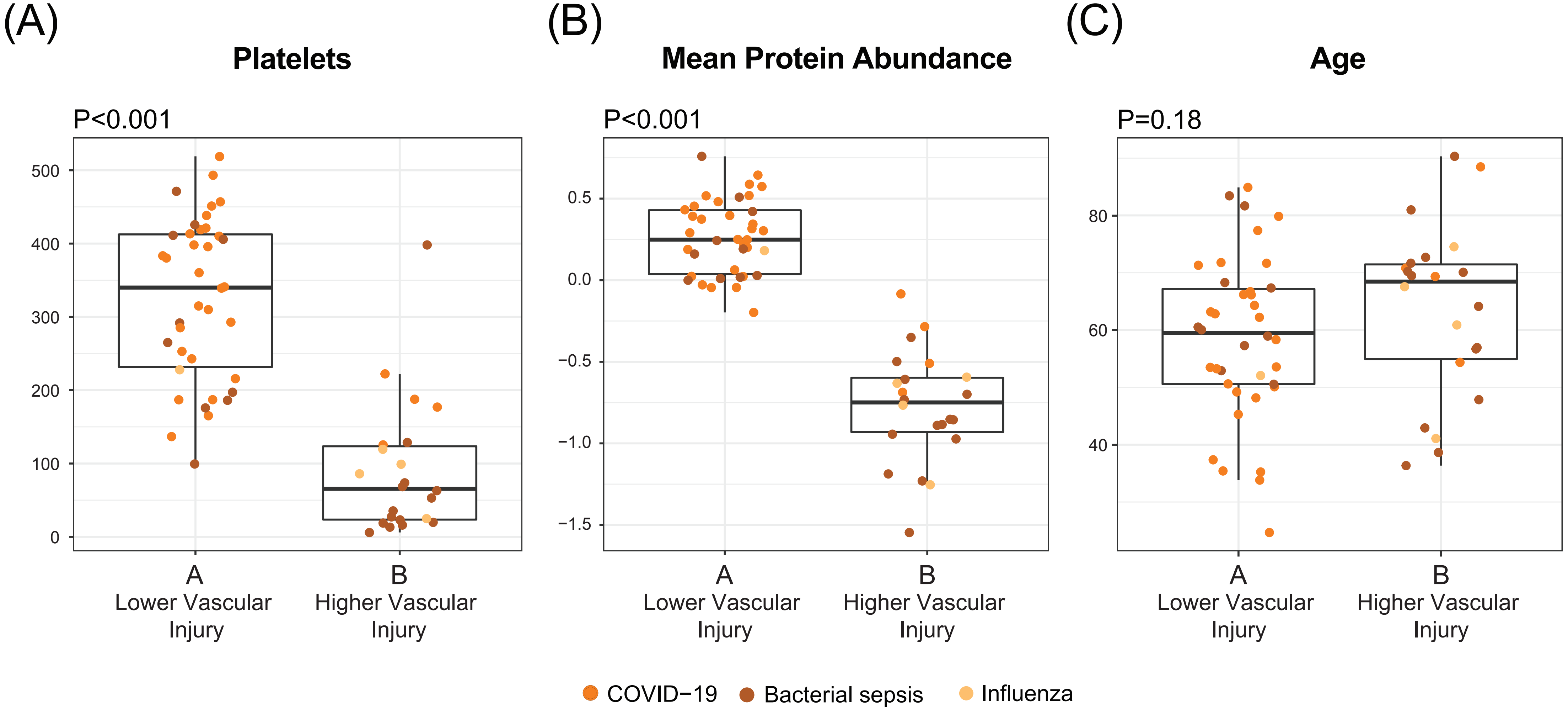


**Supplementary Figure 5: Platelet count (A), mean protein abundance (B) and age (C) by vascular injury in the two clusters defined in the ARDS cohort (N=60).** Both platelet levels and mean protein abundance are significantly lower in cluster B (P<0.001). There were no significant age differences between the two patient clusters (P=0.18). The boxes indicate the interquantile range (IQR) of the data distribution, the line in the box represents the median value and the whiskers extend for 1.5 times the value of the IQR. Dots indicate the protein level in individual patients across the different ARDS categories: COVID-19 (orange), bacterial sepsis (brown) and influenza (mustard). Differential statistic was assessed with a two-sided Mann-Whitney U test.

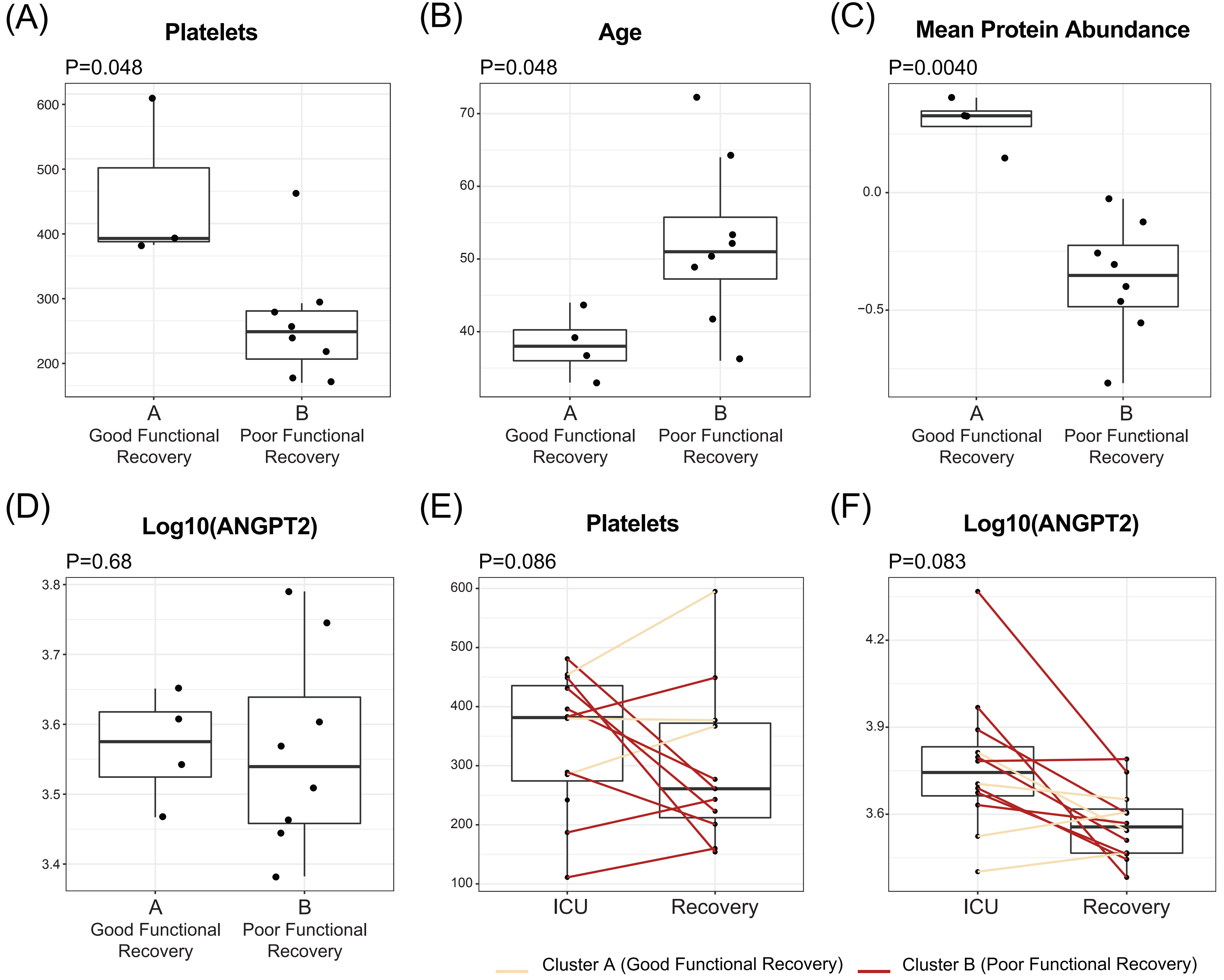


**Supplementary Figure 6: Differential analysis of clinical parameters in the recovery cohort (N=12).** (A-D) Platelet level, age, mean protein abundance and Log10(ANGPT2), in the two clusters defined in the recovery cohort, respectively. Differential statistic was assessed with a two-sided Mann-Whitney U test. (E-F) Platelet and Log10(ANGPT2) trajectory from ICU to recovery time point, respectively. The P-value refers to the comparison of the difference in protein abundance (delta) from ICU to recovery timepoint between the two functional recovery group (good versus poor functional recovery). The boxes indicate the interquantile range (IQR) of the data distribution, the line in the box represents the median value and the whiskers extend for 1.5 times the value of the IQR. Dots indicate the protein level in individual patients. Values from the same patient are linked by a line and colored according to the corresponding heatmap cluster: A (cream) or B (red). Differential statistic of the protein trajectories between the two patient clusters was computed with a linear model.

Supplementary Table 2. Baseline characteristics of the *at risk*, *ARDS,* and *recovery* cohorts by vascular signature

| | At Risk Cohort[A] | | | | ARDS Cohort[A] | | | Recovery Cohort[A] | | |
|---|---|---|---|---|---|---|---|---|---|---|
| **Characteristic** | Cluster A | Cluster B | Cluster C | P-value[C] | Cluster A | Cluster B | P-value[B] | Cluster A | Cluster B | p-value[B] |
| **N** | 29 | 21 | 9 | | 38 | 22 | | 4 | 12 | |
| **Age** | 61 (52, 67) | 77 (69,84) | 77 (72,82) | **0.002** | 59 (50,67) | 68 (55,71) | 0.2 | 39 (36,41) | 51 (47, 56) | **0.048** |
| **Sex** | | | | 0.14 | | | 0.4 | | | 0.2 |
| Female | 13 (45%) | 4 (19%) | 4 (44%) | | 10 (26%) | 8 (36%) | | 0 (0%) | 4 (50%) | |
| Male | 16 (55%) | 17 (81%) | 5 (56%) | | 28 (74%) | 14 (64%) | | 4 (100%) | 4 (50%) | |
| **Race** | | | | 0.8 | | | 0.14 | | | >0.9 |
| Asian | 2 (6.9%) | 0 (0%) | 0 (0%) | | 2 (5.3%) | 5 (23%) | | | | |
| Black | 4 (14%) | 3 (14%) | 2 (22%) | | 3 (7.9%) | 3 (14%) | | 0 (0%) | 1 (12%) | |
| Other/unknown | 12 (41%) | 6 (29%) | 3 (33%) | | 20 (53%) | 7 (32%) | | 3 (75%) | 6 (75%) | |
| White | 11 (38%) | 12 (57%) | 4 (44%) | | 13 (34%) | 7 (32%) | | 1 (25%) | 1 (12%) | |
| **Co-morbidities** | | | | | | | | | | |
| BMI | 28 (25,31) | 28 (23,33) | 23 (21,31) | 0.3 | 28 (26,34) | 25 (21, 27) | **0.012** | 30 (28,35) | 27 (26,28) | 0.2 |
| Hypertension | 12 (41%) | 13 (62%) | 6 (67%) | 0.3 | 18 (47%) | 8 (36%) | 0.4 | 0 (0%) | 4 (50%) | 0.2 |
| CAD | 3 (10%) | 5 (24%) | 3 (33%) | 0.2 | 3 (7.9%) | 4 (18%) | 0.4 | 0 (0%) | 0 (0%) | |
| CKD | 1 (3.4%) | 0 (0%) | 2 (22%) | 0.058 | 2 (5.3%) | 2 (9.1%) | 0.6 | 0 (0%) | 0 (0%) | |
| Heart Failure | 2 (6.9%) | 2 (9.5%) | 3 (33%) | 0.14 | 1 (2.6%) | 2 (9.1%) | 0.5 | 0 (0%) | 0 (0%) | |
| Cancer | 2 (6.9%) | 3 (14%) | 4 (44%) | **0.043** | 1 (2.6%) | 14 (64%) | **<0.001** | 0 (0%) | 0 (0%) | |
| **Severity of illness** | | | | | | | | | | |
| SOFA | | | | | 10 (8, 12) | 10 (9,12) | >0.9 | 10 (7,11) | 9 (8,10) | >0.9 |
| **Labs** | | | | | | | | | | |
| WBC | 4.6 (4.2,6.3) | 6.0 (3.7,7.8) | 4.4 (3.6,5.2) | 0.5 | 11 (8,16) | 8 (1,18) | 0.3 | 9.8 (8.6,10.6) | 8.2 (6.7,9.3) | 0.3 |
| Lymphocytes | 0.8 (0.6, 1.0) | 0.5 (0.3,0.9) | 0.5 (0.3,1.1) | 0.072 | 0.8 (0.6,1.4) | 0.4 (0.3,0.7) | 0.2 | 0.9 (0.7,1.1) | 0.7 (0.6,1.6) | >0.9 |
| Platelet | 177 (140,241) | 141 (100,217) | 114 (27,144) | **0.032** | 332 (244,398) | 88 (50,129) | **<0.001** | 342 (266,414) | 312 (214,409) | 0.6 |
| D-Dimer | 237 (191,389) | 868 (545,1,088) | 654 (561,1,389) | **0.031** | 979 (583,2,084) | 2,052 (572,3,540) | 0.7 | 668 (594,2,032) | 2,169 (983,2,221) | 0.4 |
| Fibrinogen | 492 (476,504) | 618 (533,686) | 390 (302,479) | 0.3 | 788 (674,951) | 543 (478 645) | **0.025** | 699 (699,699) | 550 (432,669) | >0.9 |
| aPTT | 32 (29,33) | 31 (28,33) | 34 (29,36) | 0.5 | 33 (29,39) | 34 (29,47) | 0.5 | 29 (26,31) | 30 (28,32) | 0.8 |
| PT | 13 (13,14) | 13 (12,15) | 14 (13,15) | 0.9 | 15(13,17) | 14 (13,17) | 0.8 | 13 (12,14) | 13 (12,13) | >0.9 |
| Creatinine | 0.7 (0.6,0.9) | 0.8 (0.7,1.1) | 1.5 (1.0,2.6) | **0.011** | 1.1 (0.7,1.9) | 1.7 (1.2,3.3) | **0.022** | 0.6 (0.5,0.8) | 0.6 (0.5,0.9) | >0.9 |
| **Ventilator** | | | | | | | | | | |
| P:F ratio | | | | | 94 (72,130) | 184 (107,235) | **0.010** | 78 (70,96) | 112 (96, 154) | 0.2 |
| Ventilator ratio | | | | | 1.85 (1.5,2.4) | 2.46 (2.0, 3.0) | 0.058 | 1.86 (1.63,2.00) | 1.90 (0.94, 1.94) | >0.9 |
| PEEP | | | | | 12 (9,15) | 8 (5,12) | **0.011** | 11 (10, 3) | 11(10,14) | >0.9 |
| Driving | | | | | 13.0 (11,18) | 14 (12,16) | 0.9 | 13 (12,14) | 17(13,18) | 0.7 |

[A] Statistics presented: Median (IQR); n (%). [B] Statistical tests performed: Wilcoxon rank-sum test; Fisher's exact test. [C] Statistical tests performed: Wilcoxon rank-sum test; chi-square test of independence; Fisher's exact test. BMI = body mass index, CAD = coronary artery disease, CKD = chronic kidney disease, SOFA = sequential organ failure assessment score, WBC = white blood cell count, P:F = partial pressure of arterial oxygen to fraction of inspired oxygen ratio, PEEP = positive end expiratory pressure.
